# Mandatory social distancing associated with increased doubling time: an example using hyperlocal data

**DOI:** 10.1101/2020.04.15.20067272

**Authors:** Mark H. Ebell, Grace Bagwell-Adams

## Abstract

In a hyperlocal analysis of doubling time of COVID-19, we found that a county that implemented mandatory social distancing and other measures to reduce spread of infection saw an earlier increase in doubling time than surrounding counties.

---

Mandatory social distancing has been shown in both observational and modeling studies to decrease spread of infectious diseases including SARS-CoV2. (1) It has been widely implemented in the United States during the SARS-CoV2 pandemic, but this implementation occurred at different times and with different levels of adherence. The state of Georgia has 159 relatively small counties, making it possible to see the impact of different distancing policies in neighboring counties. We present an example using this kind of hyperlocal data in the state of Georgia.

In Georgia, Clarke County was among the first counties to adopt a mandatory policy of Sheltering In Place (SIP) with two primary objectives: 1) prohibition of assemblages, events, and gatherings of more than 10 persons; and 2) requiring individuals to remain at home 24 hours a day, with very limited exceptions of essential travel. The policy was adopted on March 19 and implemented on March 20, 2020. These measures were not adopted by any surrounding counties except for one neighboring county (Oconee) that implemented a similar policy on March 24, 2020. A statewide SIP policy was not implemented until April 3, 2020. (2) This variation in SIP policies at this hyperlocal level created a natural experiment prior to the statewide policy change, allowing us to examine the relationship between SIP policy implementation and case doubling rates for Clarke (treatment county) versus surrounding counties (control group).

Statewide data regarding new cases by county have been reported since March 14, 2020. We used these data to estimate the doubling rate for Clarke County (population 127,330) and for the surrounding six counties (Jackson, Barrow, Oconee, Oglethorpe, Madison, and Walton, population 328,710).

Because of relatively small and fluctuating numbers of cases, we calculated 5 day moving averages of the percentage increase in cases in Clarke vs surrounding counties. The following formula was used to calculate doubling time, where T_d_ is doubling time and r is the 5-day moving average in the percentage increase in cases (range 0 to 100): (3)

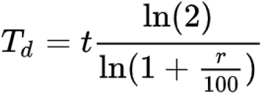

Data for new cases by date are shown in Table 1, and the doubling times for Clarke County, surrounding counties, and the state of Georgia in Figure 1. One would expect a lag of at least 5 days between implementation of mandatory distancing and an impact on disease spread, given the incubation period of the infection. A longer lag period may also be created by delays in tests results being reported, which was occurring. Assuming a 5-day median time of incubation (3) plus a 5-day lag in test results is consistent with the visual separation in doubling time curves occurring approximately 10 days after mandatory SIP went into place in Clarke County. The lag after the state SIP is likely to be shorter due to more rapid return of results as the pandemic progresses.

**Table 1.** New cases by for Clarke County, surrounding counties, and the state of Georgia.

|  | **Doubling time (days)** |  |  |
| --- | --- | --- | --- |
| **Date** | **Clarke County** | **Counties surrounding Clarke** | **State of Georgia** |
| 3/19/20 | 1.9 |  | 2.3 |
| 3/20/20 * | 2.0 |  | 2.1 |
| 3/21/20 | 2.8 | 2.1 | 2.2 |
| 3/22/20 | 2.6 | 2.6 | 2.3 |
| 3/23/20 | 3.1 | 1.9 | 2.4 |
| 3/24/20 | 4.3 | 1.6 | 2.5 |
| 3/25/20 | 5.0 | 1.9 | 3.6 |
| 3/26/20 | 3.5 | 1.7 | 3.4 |
| 3/27/20 | 2.9 | 1.7 | 2.9 |
| 3/28/20 | 3.7 | 2.2 | 3.2 |
| 3/29/20 | 4.6 | 2.7 | 3.9 |
| 3/30/20 | 4.4 | 3.5 | 4.2 |
| 3/31/20 | 4.6 | 2.7 | 3.7 |
| 4/1/20 | 7.2 | 3.0 | 4.0 |
| 4/2/20 | 7.7 | 3.0 | 4.2 |
| 4/3/20 + | 6.9 | 3.1 | 4.3 |
| 4/4/20 | 8.9 | 2.9 | 4.3 |
| 4/5/20 | 16.1 | 6.2 | 6.2 |
| 4/6/20 | 11.4 | 7.4 | 7.6 |
| 4/7/20 | 9.7 | 5.4 | 6.9 |
| 4/8/20 | 9.8 | 5.7 | 6.5 |
| 4/9/20 | 11.4 | 6.0 | 6.4 |
| 4/10/20 | 12.7 | 5.7 | 6.3 |
| 4/11/20 | 19.6 | 6.1 | 6.7 |
| 4/12/20 | 45.7 | 9.9 | 10.0 |

\* Date that SIP was mandated in Clarke County
+ Date that SIP was mandated for the state of Georgia

**Figure 1.**
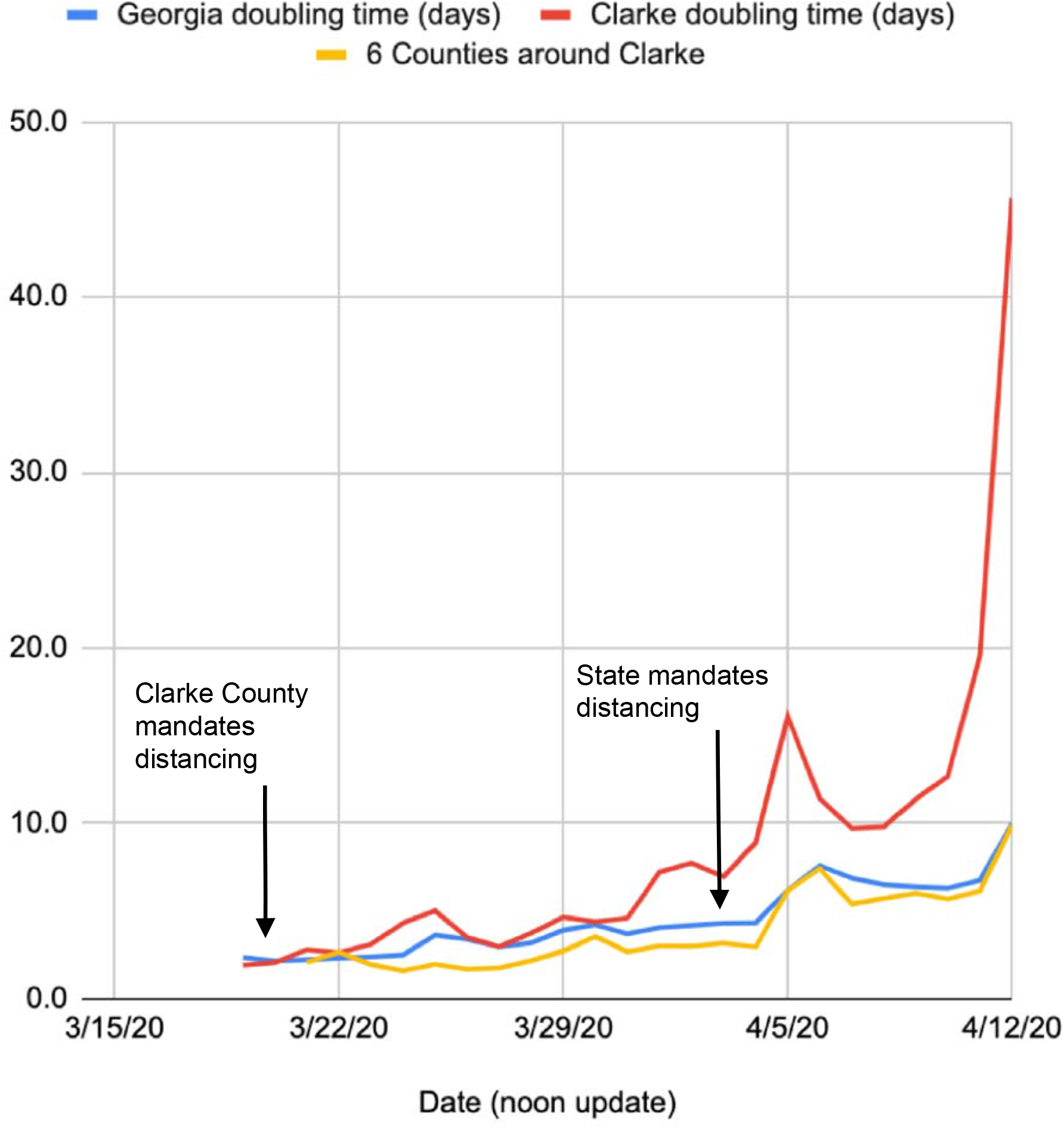
Doubling times for Clarke County, 6 surrounding counties, and the state of Georgia

Limitations include the fact that county of residence may have been incorrectly attributed. However, this would bias results against Clarke County, since it has the only hospitals in the 7-county region, meaning some patients who reside outside of Clarke may be mistakenly attributed to Clarke County. Adherence to distancing in Clarke County may have been better than in surrounding counties regardless of local mandates. However, demographic and socioeconomic characteristics, such as the unemployment rate, uninsured rate, and median income, of the region are similar across the seven-county region (with the exception of Oconee county, which has higher concentrations of wealth, education, and access to care). (4,5,6)

In conclusion, we observed a favorable impact on doubling time that corresponded to the earlier implementation of mandatory distancing in Clarke County compared to surrounding counties and the state of Georgia.

## Data Availability

Data are available on request.

## References

1. Nussbaumer-Streit B, Mayr V, Dobrescu AI, et al. Quarantine alone or in combination with other public health measures to control COVID-19: a rapid review. Cochrane Systematic Review published April 8, 2020. https://doi.org/10.1002/14651858.CD013574

2. Georgia State Policy: Issuing a statewide shelter in place to stop the spread of COVID-19. https://gov.georgia.gov/executive-action/executive-orders/2020-executive-orders. Date last reviewed 4/14/20.

3. Lauer SA, Grantz KH, Bi Q, et al. The incubation period of coronavirus disease 2019 (COVID- 19) from publicly reported confirmed cases: estimation and application.

4. Bagwell-Adams, G, Bramlett, M. Athens Wellbeing Project Health Report. https://athenswellbeingproject.org/data. Date last reviewed 4/14/20.

5. U.S. Census Bureau. 2019 Georgia County Quick Facts. https://www.census.gov/quickfacts/GA. Date last reviewed: 4/13/20.

6. Robert Wood Johnson Foundation County Health Rankings. 2019 County Health Reports. https://www.countyhealthrankings.org. Date last reviewed: 4/13/20.

